# Patients with end-stage kidney disease and COVID-19 are commonly hospitalized early during COVID-19 illness: an opportunity for early intervention

**DOI:** 10.1101/2024.01.23.24301661

**Authors:** Rituvanthikaa Seethapathy, Qiyu Wang, Nurit Katz-Agranov, Ian Strohbehn, Daiana Moreno, Destiny Harden, Roby P. Bhattacharyya, Meghan E. Sise

## Abstract

Antiviral medications such as remdesivir, molnupiravir, and nirmatrelvir/ritonavir are most effective when used early in the course of coronavirus disease 2019 (COVID-19). These medications are mainly authorized for outpatient use in high risk populations. End-stage kidney disease (ESKD) is among the strongest risk factors for mortality from COVID-19, however, therapeutic options have been lacking in this patient population given exclusion of ESKD in the registrational trials of antiviral therapy leading to limited FDA approval. In our retrospective study of patients with ESKD on dialysis admitted for symptomatic COVID-19 from March 2020 to January 2020, we found that majority of patients (>80%) were admitted to the hospital early during their disease course (within 5 days of symptom onset). Despite this pattern of early admission, there was a high risk of respiratory failure within 90 days since admission (30%) among this population. We argue that this unique pattern of early presentation and high risk of progression to respiratory failure of the ESKD patients suggests an opportunity for further research to determine if outpatient antiviral therapies should be expanded to patients with ESKD to address the huge unmet need of therapeutic intervention in this vulnerable population.

## Introduction

ESKD is among the strongest risk factors for mortality from COVID-19, even in vaccinated patients (1, 2). Antiviral medications authorized for COVID-19 provide maximal benefit in preventing progression to severe disease when used early in the course of COVID-19, however ESKD population were excluded in these registrational trials (3-5). Remdesivir, molnupiravir, and nirmatrelvir/ritonavir are antiviral therapies authorized for outpatients with COVID-19 who have mild symptoms and are at high risk of progression to severe disease. The end-stage kidney disease (ESKD) population is at high risk for severe disease and death, and may potentially derive benefit from early use of antiviral therapy. As patients with ESKD are highly linked to care and at high risk for complications, we hypothesized that this patient population is more likely to be admitted to the hospital within the early window (defined as within 5 days of symptom onset) when antiviral therapies are most effective. This pattern of admission would likely create an opportunity for clinical research and early therapeutic interventions to address the urgent unmet need of treatment options in ESKD population with COVID-19. Among patients with ESKD who presented within 5 days of symptom onset, we sought to determine the risk of respiratory failure during their disease course.

## Methods

### Study population

Using dialysis records, we identified any patient with ESKD on dialysis who was admitted to Massachusetts General Hospital (MGH) with COVID-19. Inclusion criteria included: (1) Adults age ≥18; (2) COVID-19 infection (confirmed by RT-qPCR of the nasopharyngeal or oropharyngeal specimen) diagnosed within 72 hours of admission; (3) ESKD diagnosed >90 days before index admission. Exclusion criteria included: 1) Transfer from an outside facility after diagnosis of COVID-19 (due to incomplete records of symptom and treatment history); 2) Positive RT-qPCR determined to be falsely positive or recovered COVID-19 based on MGH infection control criteria; 3) Hospital-acquired COVID-19 (diagnosed >72 hours after admission). Our institutional review board approved the protocol and waived the need for informed consent. Research was conducted in accordance with the Helsinki Declaration.

### Data collection

We manually extracted the date of symptom onset as documented in the admission note. COVID-19 associated symptoms included cough, dyspnea, fatigue, fever, rhinorrhea, chills, malaise, myalgia, loss of smell or taste, nausea, or diarrhea. Patients with ESKD were considered *asymptomatic* if they never developed symptoms attributed to COVID-19 during the hospitalization. We determined the severity of illness at hospital presentation by recording the vital signs including heart rate, blood pressure, and need for oxygen within the first 12 hours of hospital presentation. We obtained comorbidities, medications, and admission laboratory studies from the Research Patient Data Registry and determined COVID-19 vaccination status by chart review.

### Primary and secondary outcome

The primary outcome was the proportion of patients with ESKD who were admitted within 5 days of symptom onset. The secondary outcome was the risk of respiratory failure defined by invasive mechanical ventilation or death from respiratory failure within 90 days among patients who presented to the hospital within 5 days of symptom onset. We calculated time of hospital admission to development of respiratory failure among patients who developed the outcome. We performed sensitivity analysis among patients admitted during the Delta and Omicron waves in Boston, Massachusetts (July 15, 2021 – December 24, 2021 and December 25, 2021 – January 24, 2022, respectively), and among patients receiving at least one dose of an authorized COVID-19 vaccine. Because multiple patients with ESKD admitted to our center were enrolled in a placebo controlled trial of remdesivir during the study period, and nirmatrelvir/ritonavir and molnupiravir were not yet authorized for use in the study period, we did not evaluate the effect of COVID-19 directed therapies on treatment outcomes.

## Results

A total of 124 patients with ESKD were admitted to MGH with COVID-19 between March 21, 2020 - January 24, 2022. After applying the exclusion criteria shown in **Figure 1**, 99 patients with community-acquired COVID-19 were included. Thirty patients (30%) remained asymptomatic throughout their entire hospitalization. Among the 69 (70%) who were symptomatic, 56 (81%) were admitted within 5 days of symptom onset and 13 (19%) were admitted more than 5 days after symptom onset (**Figure 1**). The median time from symptom onset to hospitalization was 2 days (interquatile range [IQR] 1—4 days).

**Figure 1.**
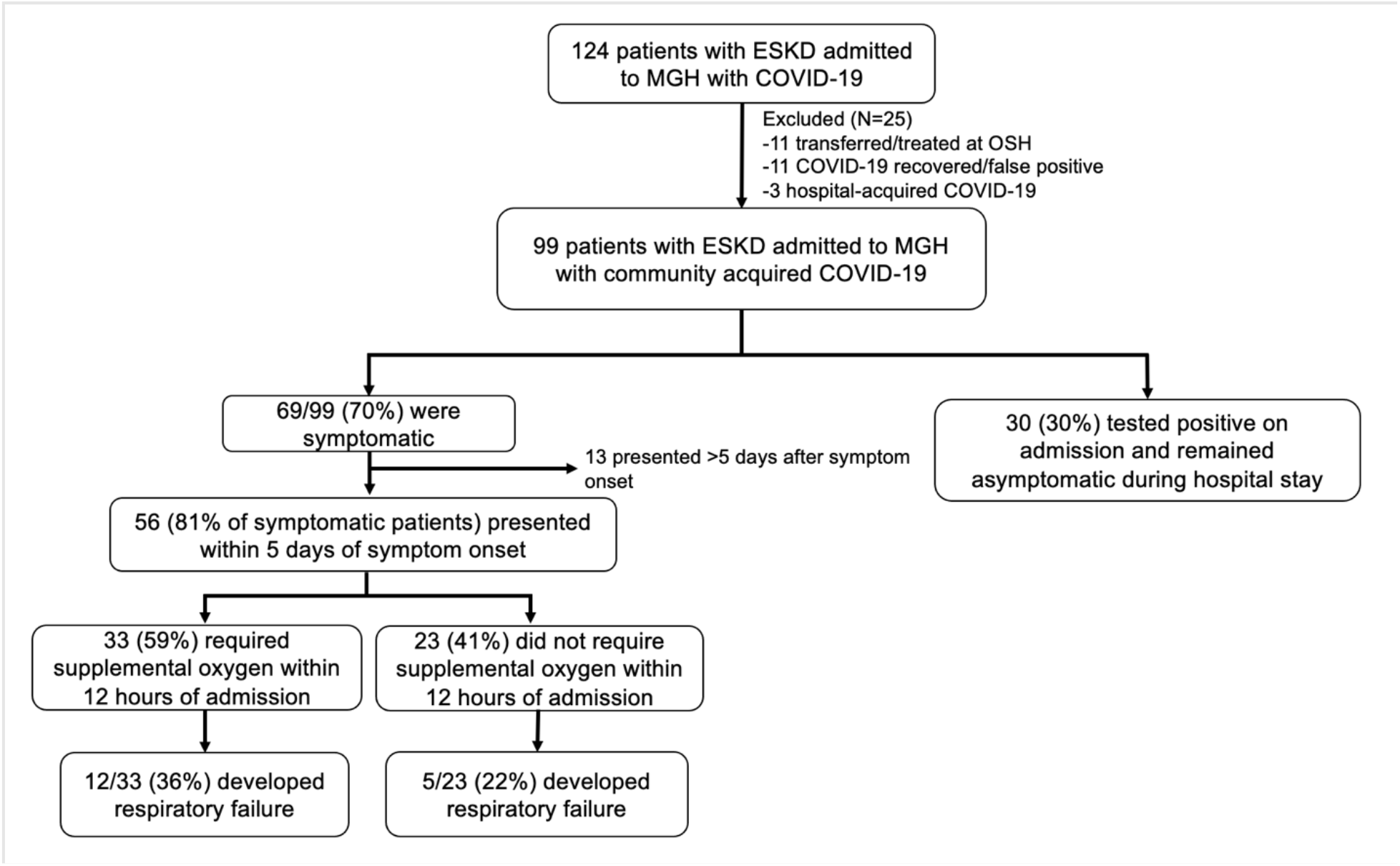
Patient flow. Abbreviations: ESKD = end stage kidney disease, MGH = Massachusetts General Hospital, COVID-19 = coronavirus disease 2019, OSH = outside hospital. Among the 17 patients with respiratory failure, 11 died within 90 days.

Among the 56 patients who presented within 5 days of symptom onset, 33 out of 56 (59%) required supplemental oxygen within 12 hours of admission, and 12 out of those 33 (36%) subsequently developed respiratory failure and 9 died of respiratory failure within 90 days. Among those who did not require oxygen within 12 hours of admission, 5 out of 23 (17%) progressed to respiratory failure and 2 died of respiratory failure within 90 days (**Figure 1**). The median time from hospital admission to respiratory failure was 23 days (IQR 9—44 days).

When limiting our analysis to the Delta (N=9) and Omicron (N=8) waves in Boston and to only vaccinated patients (N=25), >90% presented within 5 days of symptom onset and >30% developed respiratory failure.

## Discussion

We found that the vast majority of patients with ESKD who were admitted for COVID-19 presented to the hospital within 5 days of symptom onset, during the window of maximal expected efficacy of antiviral therapy. Despite early presentation, these patients had a high risk of respiratory failure, even among the subset who did not require oxygen during the first 12 hours of admission. This finding was consistent in the Delta and Omicron waves in Boston, MA, and regardless of vaccination status. Early hospitalization among ESKD differs from the general population, in which hospitalization typically occurs in the second week of illness, rendering antiviral therapy less efficacious(6, 7). This pattern of early presentation is likely because ESKD population is highly linked to care, is screened for COVID-19 symptoms three times per week, and has high rates of referral for COVID-19 testing among symptomatic patients.

Our study confirms prior larger studies showing that patients with ESKD on dialysis have a high risk of respiratory failure and death from COVID-19 even after vaccination. Bell and colleagues detected a 7% one-month mortality among fully vaccinated patients with ESKD receiving dialysis who developed breakthrough COVID-19 infection in a large population study(8). This ongoing high risk of respiratory failure and death makes patients with ESKD high priority candidates for early antiviral therapy.

A summary of currently available outpatient therapies for COVID-19 is shown in **Table 1**. . However, ESKD patients are not among the main beneficiaries given their early hospital presentation in the disease course making them ineligible for outpatient therapies (only remdesivir is approved for inpatient use) and exclusion due to ESKD status. Nirmatrelvir/ ritonavir, a protease inhibitor, received EUA for use in patients within 5 days of symptom onset after a large trial showed it reduced the risk of hospitalization or death by 89%; however, nirmaltervir/ritonavir is renally eliminated and not recommended in patients with eGFR < 30mL/min/1.73m^2^; dose adjustment is required for patients with eGFR 30-59mL/min/1.73m^2^ (5). Molnupiravir, a nucleoside analog that inhibits viral replication through lethal mutagenesis, led to a 30% reduction in hospitalization or death among outpatients at risk for hospitalization who were treated within 5 days (4). Although patients with ESKD were excluded from trials of molnupiravir, its EUA does not exclude patients based on eGFR. The approval for remdesivir was recently expanded to high-risk outpatients with mild-to-moderate COVID-19 after a trial showed that a three-day course of remdesivir given within 7 days of symptom onset lowered the risk of hospitalization by 87% (3). Although remdesivir is not currently approved for patients with ESKD or on dialysis, prior single and multi-center case series of off-label remdesivir use in patients with ESKD have suggested that it is safe and well-tolerated (9-14). Definitive safety and efficacy data will hopefully follow from the “Study to Evaluate the Efficacy and Safety of Remdesivir in Participants with Severely Reduced Kidney Function Who Are Hospitalized for Coronavirus Disease 2019” (REDPINE Trial), a randomized controlled trial recruiting patients with severe COVID-19 who have eGFR <30 mL/min/1.73m^2^ or are on dialysis (NCT04745351). Importantly, remdesivir had smaller effect sizes in trials limited to inpatient use but showed striking benefits for outpatients, which is generally attributed to its use earlier during the course of illness (3, 15, 16). Since patients with ESKD are frequently admitted within the first 5 days of symptoms, our findings may inform the risk-benefit analysis in this population that was excluded from the original trials. Furthermore, while outpatient remdesivir is challenging due to the need for parenteral access, it is feasible to administer to admitted patients who present early in the disease course when antivirals are most effective. Finally, monoclonal antibodies (mAbs) are metabolized by target-mediated elimination and thus are safe and approved for all levels of kidney function, and are highly effective at preventing hospitalization when given early and when appropriately matched with the infecting variant (17). However, inpatient mAb use is generally restricted to emergency investigational new drug applications, greatly limiting its use. Since the vast majority of ESKD patients are admitted early in their illness, their window for outpatient mAb use is limited.

**Table 1.** Current available outpatient antiviral therapies for COVID-19. Abbreviations: COVID-19 = coronavirus disease 2019, ESKD = end-stage kidney disease, IV = intravenous, eGFR = estimated glomerular filtration rate, EUA = emergency use authorization, IM = intramuscular

| Therapy | Effect size | Data/Approval in ESKD | Downside |
| --- | --- | --- | --- |
| **Remdesivir** | Reduced hospitalization or death by 87% in high-risk outpatients | Not approved for eGFR < 30 ml/min due to renal elimination of active metabolite and cyclodextrin carrier, but used in numerous off-label studies in ESKD | IV administration x 3 days |
| **Molnupiravir** | Reduced hospitalization or death by 30% in high-risk outpatients | Not studied in eGFR < 30 ml/min, but EUA includes all levels of eGFR | Limited effect size |
| **Nirmatrelvir/ritonavir** | Reduced hospitalization or death by 89% in high-risk outpatients | Renally eliminated, dose reductions for eGFR 30-59 ml/min, Not studied or authorized for eGFR < 30 ml/min. | Numerous drug-drug interactions (ritonavir) |
| **Monoclonal Antibodies (Bebtelovimab current)** | Varies across studies (80-90% reduction in hospitalization) | Metabolism unlikely to be affected by kidney function | IV/IM administration; highly susceptible to loss of activity against variants and subvariants |

Our study is limited by its retrospective design and the fact that it is derived from a single center limiting its generalizability. Second, most patients included in the analysis were admitted during the first wave of COVID-19, and number of admissions for patients with ESKD during the delta and omicron waves were lower. Although COVID-19 outcomes have improved over time, likely due to increasing population immunity through vaccination and prior infection, mortality data in patients with ESKD receiving dialysis remain limited in the Omicron era. In our study, we found that among the small number of patients that did require hospitalization, the risk of respiratory failure and death remained high. Future studies are needed to evaluate the mortality risk among patients with ESKD diagnosed with COVID-19 in both inpatient and outpatient settings in the Omicron era, as our study only included patients sick enough to warrant hospitalization.

In conclusion, we found that >80% of patients with ESKD on dialysis admitted for symptomatic COVID-19 presented within 5 days of symptom onset (median 2 days after symptom onset IQR 1-4). We speculate that because of this, inpatient antiviral therapies may be more effective in the ESKD population than in a typical inpatient population with COVID-19 that presents later in the disease course. We also detected a high risk of respiratory failure in patients with ESKD, highlighting the important unmet needs for early treatment strategies in this population. Their unique pattern of early presentation and high risk of progression to respiratory failure of the ESKD patients suggests an opportunity when clinical research of antiviral efficacy can be taken place and indication for outpatient use of antiviral therapies to be expanded to address the huge unmet need of therapeutic intervention in this vulnerable population.

## Data Availability

De-identified data is available upon reasonable request to Dr. Meghan Sise via email and after execution of a data use agreement with Mass General Brigham.

## Acknowledgements

This work was funded by Gilead through an investigator-initiated grant to Massachusetts General Hospital (MGH) awarded to Dr. Sise. All aspects of the study (protocol design, data capture, analysis, and manuscript preparation) were performed by Dr. Sise and her co-authors. RS, NK, DM, DH performed chart review and data collection. QW, MS designed study, performed data analysis and manuscript drafting. IS captured and organized clinical data. RB critically revised the manuscript. All authors participated in manuscript revision and are in agreement with the content of the manuscript.

## Financial Disclosures

**Dr. Sise:** Research funding: Angion, Otsuka, Gilead, Abbvie, Cabaletta, Novartis, EMD-Serono Scientific advisory boards: Vera, Travere, Calliditas, Mallinckrodt, Novartis

DSMB member: Alpine Immunosciences

Consulting: Resonance

All remaining authors have nothing to disclose.

## References

1. Chronic kidney disease is a key risk factor for severe COVID-19: a call to action by the ERA-EDTA. Nephrol Dial Transplant 2021;36:87–94.

2. Bell S, Campbell J, Lambourg E, Watters C, O’Neil M, Almond A, Buck K, et al. The Impact of Vaccination on Incidence and Outcomes of SARS-CoV-2 Infection in Patients with Kidney Failure in Scotland. Journal of the American Society of Nephrology 2022:ASN.2022010046.

3. Gottlieb RL, Vaca CE, Paredes R, Mera J, Webb BJ, Perez G, Oguchi G, et al. Early Remdesivir to Prevent Progression to Severe Covid-19 in Outpatients. N Engl J Med 2022;386:305–315.

4. Jayk Bernal A, Gomes da Silva MM, Musungaie DB, Kovalchuk E, Gonzalez A, Delos Reyes V, Martin-Quiros A, et al. Molnupiravir for Oral Treatment of Covid-19 in Nonhospitalized Patients. N Engl J Med 2022;386:509–520.

5. Hammond J, Leister-Tebbe H, Gardner A, Abreu P, Bao W, Wisemandle W, Baniecki M, et al. Oral Nirmatrelvir for High-Risk, Nonhospitalized Adults with Covid-19. N Engl J Med 2022.

6. Berenguer J, Borobia AM, Ryan P, Rodriguez-Bano J, Bellon JM, Jarrin I, Carratala J, et al. Development and validation of a prediction model for 30-day mortality in hospitalised patients with COVID-19: the COVID-19 SEIMC score. Thorax 2021;76:920–929.

7. Gupta S, Hayek SS, Wang W, Chan L, Mathews KS, Melamed ML, Brenner SK, et al. Factors Associated With Death in Critically Ill Patients With Coronavirus Disease 2019 in the US. JAMA Intern Med 2020;180:1436–1447.

8. Bell S, Campbell J, Lambourg E, Watters C, O’Neil M, Almond A, Buck K, et al. The Impact of Vaccination on Incidence and Outcomes of SARS-CoV-2 Infection in Patients with Kidney Failure in Scotland. J Am Soc Nephrol 2022;33:677–686.

9. Thakare S, Gandhi C, Modi T, Bose S, Deb S, Saxena N, Katyal A, et al. Safety of Remdesivir in Patients With Acute Kidney Injury or CKD. Kidney Int Rep 2021;6:206–210.

10. Pettit NN, Pisano J, Nguyen CT, Lew AK, Hazra A, Sherer R, Mullane KM. Remdesivir Use in the Setting of Severe Renal Impairment: A Theoretical Concern or Real Risk? Clin Infect Dis 2021;73:e3990–e3995.

11. Ackley TW, McManus D, Topal JE, Cicali B, Shah S. A Valid Warning or Clinical Lore: an Evaluation of Safety Outcomes of Remdesivir in Patients with Impaired Renal Function from a Multicenter Matched Cohort. Antimicrob Agents Chemother 2021;65.

12. Aiswarya D, Arumugam V, Dineshkumar T, Gopalakrishnan N, Lamech TM, Nithya G, Sastry B, et al. Use of Remdesivir in Patients With COVID-19 on Hemodialysis: A Study of Safety and Tolerance. Kidney Int Rep 2021;6:586–593.

13. Estiverne C, Strohbehn IA, Mithani Z, Hirsch JS, Wanchoo R, Goyal PG, Lee Dryden-Peterson S, et al. Remdesivir in Patients With Estimated GFR <30 ml/min per 1.73 m(2) or on Renal Replacement Therapy. Kidney Int Rep 2021;6:835–838.

14. Seethapathy R, Zhao S, Long JD, Strohbehn IA, Sise ME. A propensity-score matched observational study of remdesivir in patients with COVID-19 and severe kidney disease. Kidney360 2021;2.

15. Young B, Tan TT, Leo YS. The place for remdesivir in COVID-19 treatment. Lancet Infect Dis 2021;21:20–21.

16. Hussain Alsayed HA, Saheb Sharif-Askari F, Saheb Sharif-Askari N, Hussain AAS, Hamid Q, Halwani R. Early administration of remdesivir to COVID-19 patients associates with higher recovery rate and lower need for ICU admission: A retrospective cohort study. PLoS One 2021;16:e0258643.

17. Focosi D, McConnell S, Casadevall A, Cappello E, Valdiserra G, Tuccori M. Monoclonal antibody therapies against SARS-CoV-2. Lancet Infect Dis 2022;22:e311–e326.

